# Intravenous immunoglobulin has epigenetic, ribosomal, and immune effects in Paediatric Acute-Onset Neuropsychiatric Syndrome

**DOI:** 10.1101/2025.03.27.25324808

**Authors:** Velda X Han, Hiroya Nishida, Brooke A Keating, Brian S Gloss, Xianzhong Lau, Ruwani Dissanayake, Jessica Hayes, Shekeeb S Mohammad, Shrujna Patel, Russell C Dale

**Affiliations:** Kids Neuroscience Centre, The Children’s Hospital at Westmead, Faculty of Medicine and Health, University of Sydney, NSW, Australia; Khoo Teck Puat-National University Children’s Medical Institute, National University Health System, Singapore, Singapore; Department of Paediatrics, Yong Loo Lin School of Medicine, National University of Singapore, Singapore; Westmead Research Hub, Westmead Institute for Medical Research, Westmead, NSW, Australia; Australian Genome Research Facility Ltd, Melbourne, VIC, Australia; Australian Genome Research Facility Ltd, Westmead, NSW, Australia; The Children’s Hospital at Westmead Clinical School, Faculty of Medicine and Health, University of Sydney, Sydney, NSW, Australia; The University of Sydney, School of Medical Sciences and Discipline of Child and Adolescent Health, Faculty of Medicine and Health, Sydney, NSW, Australia

**Keywords:** epigenetics, immune, neurodevelopmental disorders, transcriptomics, RNA sequencing

## Abstract

Paediatric acute-onset neuropsychiatric syndrome (PANS) is characterised by infection-provoked abrupt and dramatic onset of obsessive compulsive disorder (OCD) or eating restriction, along with neurodevelopmental regression. Although the aetiology of PANS remains uncertain, due to a prevailing neuroimmune hypothesis some children receive the immune modulator intravenous immunoglobulin (IVIg). Using single-cell RNA sequencing of peripheral immune cells, we examined gene expression in five children with PANS (mean age 10.2 years, range 5–17, 60% male) who were receiving open-label IVIg. Maternal autoimmunity (n=3) and familial neurodevelopmental/neuropsychiatric disorders (n=5) were present. The index PANS event occurred age 1.8–13 years, characterised by abrupt eating restriction (n=5), developmental regression (n=4), and OCD (n=3).

Single-cell RNA sequencing (144,470 cells) was performed in five patients pre- and post-IVIg, and four controls (mean age 13.5 years, range 11–16, 50% male). In PANS pre-IVIg compared to neurotypical controls, there were downregulated immune pathways (defense response, innate immunity, secretory granules) in most cell types. However, NK cells exhibited upregulated immune pathways (response to corticosteroid), supporting a baseline ‘immune dysregulation’. In PANS pre-IVIg compared to controls, ribosomal pathways were upregulated in neutrophils and CD8 T cells, but downregulated in NK cells. Post-IVIg, the previously downregulated immune pathways were upregulated in most cell types, and the baseline ribosomal pathway abnormalities were reversed. Additionally, histone modification pathways (histone methyltransferase, chromatin) were downregulated in neutrophils and NK cells post-IVIg. We propose PANS is an epigenetic immune-brain disorder with cellular epigenetic, ribosomal, and immune dysregulation. Therefore, epigenetic and immune-modulating therapies, such as IVIg, may have a critical role in treating this disabling disorder.

**Highlights:**

- Five children with PANS receiving monthly IVIg who had clinical benefit were studied
- Three young boys had a regressive (autistic) phenotype, whereas the older two girls had OCD
- Single-cell RNA sequencing showed epigenetic, ribosomal, and immune dysregulation in PANS
- IVIg treatment had epigenetic and immune-modulating effects in PANS
- IVIg altered gene expression of histone methyltransferase and chromatin genes

## Introduction

Paediatric acute-onset neuropsychiatric syndrome (PANS) is characterised by acute-onset obsessive compulsive disorder (OCD) or eating restriction, plus separation anxiety, developmental regression, and somatic symptoms such as sleep and urinary disturbances(1). PANS is classically triggered by infections, but non-infectious stress triggers have also been reported. PANS is a debilitating condition with significant functional impairments, impacting school performance and social interactions. The behavioural and psychiatric symptoms associated with PANS can be resistant to conventional psychiatric medications and psychological therapies. The exact immune mechanisms driving PANS remain poorly understood, and whether PANS is a discrete entity or part of the neurodevelopmental continuum is unclear(1). We hypothesize that PANS represents a clinical phenotype demonstrating an interplay between environmental and genetic factors, where infection or stress triggers disrupted peripheral-central immune and brain function, resulting in sudden deterioration in behaviour.

Due to the leading hypothesis that PANS is driven by abnormal immune function, immunotherapy such as steroids, antibiotics, and intravenous immunoglobulin (IVIg) have been used with varying degrees of success(2). IVIg has been reported to be beneficial in some children with severe PANS, improving psychiatric and behavioural symptoms, although randomised controlled trials have yielded mixed results, and controversy regarding the aetiology and therapeutics of PANS remain(3, 4). We previously explored gene regulation through bulk RNA sequencing in children with PANS and unexplained neurodevelopmental regression, which showed an epigenetic, immune, and ribosomal signature(5). Here, we explore the effects of IVIg on peripheral immune cells in PANS through the use of single-cell RNA sequencing.

## Materials and Methods

### Participant selection

Children (<18 years of age) met criteria for paediatric acute-onset neuropsychiatric syndrome (PANS) based on 2013 PANS diagnostic criterion(1):

- an abrupt, dramatic onset (defined as within 48 hours) of OCD or severely restricted food intake
- concurrent presence of at least two of the seven categories

`o` anxiety
`o` emotional lability and/or depression
`o` irritability, aggression, and/or oppositional behaviours
`o` behavioural regression
`o` deterioration in school performance
`o` sensory or motor abnormalities
`o` somatic signs and symptoms, including sleep disturbances, enuresis or urinary frequency.

Recruited patients had ongoing debilitating neuropsychiatric symptoms at the time of treatment, despite optimization of conventional psychological or psychiatric treatments. Recruited patients were about to commence for first time (n=2), or were receiving ongoing (n=3) open-label IVIg treatment (1.5-2g/kg per month). In Australia, PANS is currently a Medicare-approved indication for IVIg.

### Control selection

We recruited age- and sex-matched healthy children of hospital workers. The inclusion criterion for controls was absence of neurodevelopmental or neuropsychiatric disorder, autoimmune diseases, and severe allergic conditions.

### Sample collection

In children with PANS, the baseline blood sample was taken during cannulation, and the post-IVIg blood sample was taken at the end of the IVIg infusion (24 hours after infusion commencement). Both patients and controls had absence of any infections or hospitalizations a month prior to blood taking.

HIVE™ *Single-cell RNA sequencing, bioinformatic and enrichment analyses*

Single-cell RNA sequencing, using the HIVE™ platform, described in Supplementary Material, on five patients with PANS before and after IVIg treatment, as well as four age matched healthy controls.

RNA sequencing data was analyzed in the R statistical environment with tidyverse, described in Supplementary Material. For HIVE™ *single-cell RNA sequencing,* the Seurat package was used for analysis. Pathway enrichment analysis was performed via Gene Set Enrichment Analysis (GSEA) to obtain Gene Ontology (GO) pathways (FDR <0.05) using the *clusterProfiler* package.

## Ethics Approval

Ethical approval was granted by the Sydney Children’s Hospitals Network Human Research Ethics Committee (HREC/18/SCHN/227, 2021/ETH00356).

## Results

### Clinical data

Five children (mean age 10.2 (5–17) years, 60% males) diagnosed with PANS, and who were commencing IVIg (2g/kg), or were on regular IVIg treatment (1.5g/kg), were recruited. 3 out of 5 children (the males with autism) had negative trio whole exome sequencing. This is a biological study to explore biological mechanisms of therapy, rather than a study to explore clinical therapeutic effect (case histories in Supplementary table 1 and Supplementary material).

The age of the index PANS event was 1.8-13 years. Three mothers had autoimmunity (coeliac disease, hypothyroidism, ulcerative colitis), and five probands had a first-degree family history of neurodevelopmental or neuropsychiatric disease (Supplementary table). At first PANS event, four children were typically developing, one had ADHD. Four patients had preceding infections at PANS onset, but no clear trigger in one patient. In the first PANS event, symptoms included abrupt onset eating restriction (n=5), developmental regression (n=4), OCD (n=3), loss of continence (n=4), sleep disturbance (n=2), and emotional dysregulation (n=3). All five had one or more further events, and four have had multiple events, almost all triggered by infection. Abrupt worsening of OCD has been the dominant feature in the subsequent events. Five have received psychological therapies, and four have had conventional medical therapies with some benefit. Convincing clinical benefit of IVIg was reported in 4 of the 5 patients (only partial benefit in other patient), but headache was a common side effect in all patients. All five patients currently carry ongoing emotional and OCD diagnoses, and the three boys have ASD diagnoses (Supplementary material).

*Single-Cell RNA Sequencing (scRNAseq): UMAP and differentially expressed genes* A total of 144,470 cells were sequenced across 14 samples from five patients pre- and post-IVIg treatment (mean age 10.2, range 5-17 years, 60% males), and four controls, mean age 13.5 (11–16) years, 50% males). Uniform manifold approximation and projection analysis of biological samples revealed 11 distinct cell clusters (Figure 1A). Differentially expressed genes (DEGs) with FDR <0.05 of the seven cell types with most DEGs, ranged from 49 to 947 DEGs per cell type in pre-IVIg vs control comparison (Figure 1B, left), with the largest DEGs in neutrophils and B cells. There were 109 to 2347 DEGs per cell type in post vs pre-IVIg comparison, predominantly in CD8 T cells, followed by non-classical monocytes and neutrophils (Figure 1B, right).

**Figure 1:**
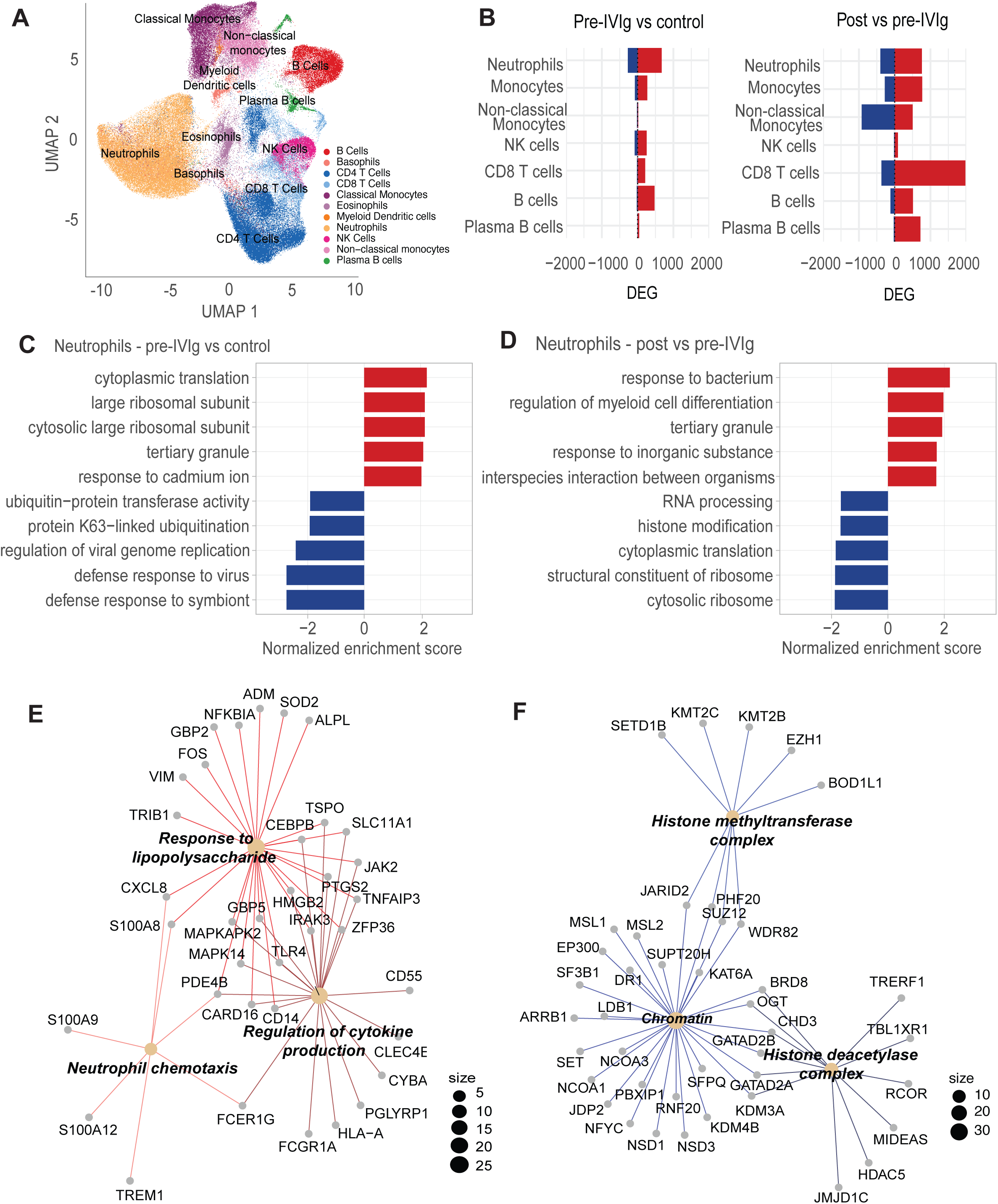
**Single cell-RNA sequencing in children with PANS pre and post IVIg** (A) Uniform manifold approximation and projection (UMAP) analysis identified 11 distinct cell clusters (B) Bar chart of differentially expressed genes (DEGs) across 7 cell types in PANS pre-IVIg vs control (left), and PANS post- vs pre-IVIg (right), showing the number of upregulated (red) or downregulated (blue) DEGs. (C) **Neutrophils-PANS pre-IVIg vs control comparison**: Bar plot of the top 5 upregulated and downregulated GSEA GO pathways (D) **Neutrophils-PANS post- vs pre-IVIg comparison:** Bar plot of the top 5 upregulated and downregulated GSEA GO pathways (E) Connectivity network enrichment plot of the upregulated ‘response to bacterium’ GO pathway in Neutrophils (PANS post- vs pre-IVIg comparison). Dot size corresponds to number of genes enriching that pathway. (F) Connectivity network enrichment plot of the downregulated ‘histone modification GO pathway in neutrophils (PANS post- vs pre-IVIg comparison). Dot size corresponds to number of genes enriching that pathway.

GSEA analysis used a ranked gene list derived enriched GO pathways (FDR <0.05) for individual cell types. We focused on results in neutrophils and NK cells, while the results for other cell types are in Supplementary Materials.

### Neutrophils

#### PANS pre-IVIg vs control comparison

In PANS pre-IVIg vs control comparison, top five upregulated GO pathways in neutrophils included ribosomal/translation pathways (Figure 1C, in red). Top five downregulated GO pathways included immune pathways such as defense response to symbiont (Figure 1C, In blue).

#### PANS post- vs pre-IVIg comparison

In PANS post- vs pre-IVIg comparison, top five upregulated GO pathways in neutrophils included immune pathways such as response to bacterium (Figure 1D, in red). We focused on the genes from the ‘response to bacterium’ pathway. A CNET plot of these genes (Figure 1E) revealed GO biological process subclusters including response to lipopolysaccharide (enriched by *TLR4, IRAK3, MAPK14, MAPKAPK2, CXCL8, TNFAIP3*), regulation of cytokine production (enriched by *CD14, JAK2*, *HMGB2, FCGR1A, FCER1G*), and neutrophil chemotaxis (enriched by *S100A8, S100A9, S100A12*).

Top five downregulated GO pathways included ribosomal/translation pathways, RNA processing, and histone modification (Figure 1D, in blue). We focused on the genes from the ‘histone modification’ pathway. A CNET plot of these genes (Figure 1F) revealed GO cellular component subclusters including histone methyltransferase (enriched by *KMT2B, KMT2C, SETD1B*), histone deacetylase complex (enriched by *HDAC5, JMJD1C*), and chromatin (enriched by *KDM3A, KDM4B, CHD3, EP300, KAT6A, GATAD2A, GATAD2B*).

### Natural Killer (NK) Cells

#### PANS pre-IVIg vs control comparison

In PANS pre-IVIg vs control comparison, top five upregulated GO pathways in NK cells included immune pathways such as response to corticosteroid and myeloid cell activation (Figure 2A, in red). Top five downregulated GO pathways included ribosomal/translation pathways (Figure 2A, In blue).

**Figure 2:**
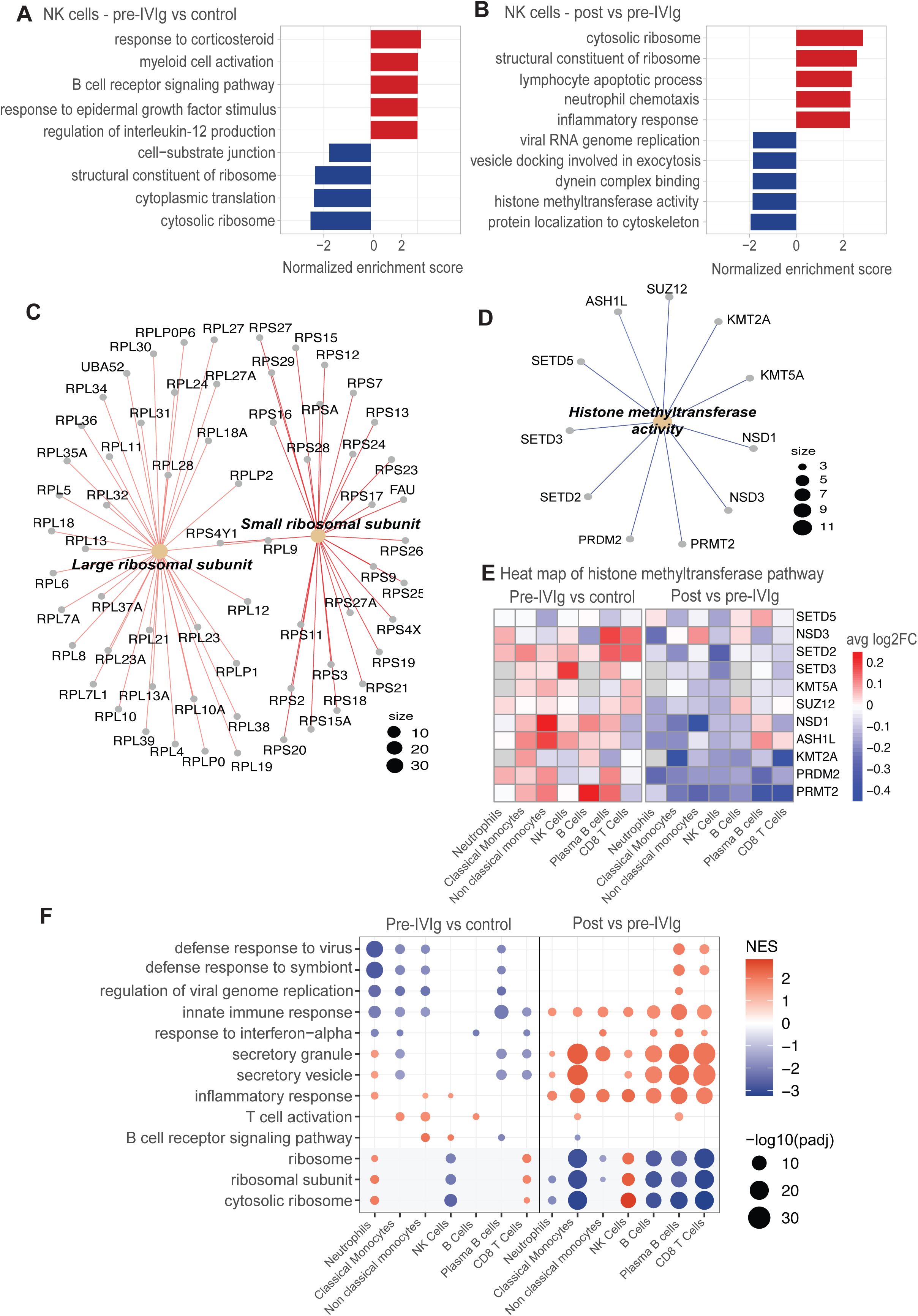
Single cell-RNA sequencing in children with PANS pre and post IVIg (A) **NK cells-PANS pre-IVIg vs control comparison**: Bar plot of top 5 upregulated and downregulated GSEA GO pathways. (B) **NK cells-PANS post- vs pre-IVIg comparison:** Bar plot of top 5 upregulated and downregulated GSEA GO pathways. (C) Connectivity network enrichment plot of the upregulated ‘cytosolic ribosome’ GO pathway in NK cells (PANS post- vs pre-IVIg) comparison. Dot size corresponds to number of genes enriching that pathway. (D) Connectivity network enrichment plot of the ‘histone methyltransferase’ GO pathway in NK cells (PANS post- vs pre-IVIg) comparison. Dot size corresponds to number of genes enriching that pathway. (E) Heatmap of genes in ‘histone methyltransferase’ GO pathway by average log2 fold change, with pre-IVIg vs control (left) and post- vs pre-IVIg comparison (right). (F) Dot plot visualising the top 5 upregulated and downregulated GSEA GO pathways across cell types in PANS pre-IVIg vs controls (left) and PANS post- vs pre-IVIg (right). Significant pathways (FDR <0.05) were simplified, and only those that were present in > 3 cell types in PANS pre-IVIg vs control comparison were plotted. Dot size represents the -log10(padj) of the pathway, while the colour intensity represents the normalized enrichment score (NES).

#### PANS post- vs pre-IVIg comparison

In PANS post vs pre-IVIg comparison, top five upregulated GO pathways in NK cells included ribosomal/translation, as well as immune pathways such as neutrophil chemotaxis, inflammatory response (Figure 2B, in red). We focused on the genes from the ‘cytosolic ribosome’ pathway. A CNET plot of these genes (Figure 2C) revealed GO cellular component subclusters large ribosomal subunit (enriched by *RPL genes*), and small ribosomal subunit (enriched by *RPS* genes).

Top five downregulated GO pathways included cytoskeletal and histone methyltransferase pathways (Figure 2B, in blue). A CNET plot of the genes in ‘histone methyltransferase’ pathway (Figure 2D) revealed enrichment of genes including *KMT2A, KMT5A, SETD2, SETD3, SETD5, ASH1L.* A heatmap of the expression of 11 genes in the ‘histone methyltransferase’ pathway showed overall upregulation in pre-IVIg PANS vs controls (Figure 2E, in red), followed by downregulation post vs pre-IVIg (Figure 2E, in blue).

### Enriched pathways across all cell types

The top five upregulated and downregulated GSEA GO pathways of individual cell types in PANS pre-IVIg vs control (Figure 2F, left) and PANS post- vs pre-IVIg (Figure 2F, right) were plotted, showing similarities and differences in pathways across cell types. In the PANS pre-IVIg vs control comparison, there were predominantly downregulated immune pathways such as defense response to virus and secretory granule/vesicle; these pathways were generally upregulated post-IVIg treatment across most cell types. In the PANS pre-IVIg vs control comparison, ribosomal pathways demonstrated diverse expression patterns across cell types, with upregulation in neutrophils and CD8 T cells, but downregulation in NK cells. These ribosomal pathways showed a reversal in direction following IVIg treatment.

## Discussion

In this single-cell RNA sequencing study of children with PANS and healthy controls, we identified key baseline alterations in epigenetic, ribosomal, and immune pathways in PANS patients, that were reversed after IVIg. In this study, the children with PANS all had debilitating behavioural symptoms and high social and academic impairments. As per previous reports of PANS, there were high rates of neurodevelopmental and neuropsychiatric disorders in family members, and maternal autoimmunity(6). We believe this supports our model of combined genetic (neurodevelopmental and immune), and environmental (maternal autoimmunity during pregnancy) factors, resulting in epigenetic immune-brain dysregulation(7).

The PANS phenotype of the three younger males included regression (autistic) features, whereas the two older females had predominant OCD symptoms. This suggests that the clinical presentation of PANS may be linked to gender and age-related vulnerabilities in neurodevelopmental processes.

This study was not a clinical trial, and PANS patients received open-label IVIg treatment. Previous randomised controlled trials of IVIg in PANS have yielded mixed results(3, 4). One trial which failed to show superiority of IVIg over placebo, involved evaluation 6 weeks after IVIg (3). In our experience, IVIg treatment improved psychiatric and behavioural symptoms, leading to better overall function, but the benefit typically lasted only 2-3 weeks. Patients often suffered further recurrent infection provoked events, and three required prolonged monthly IVIg use. This suggests that the effect of IVIg was not sustained, highlighting the need for further research to understand the baseline abnormality in PANS, in order to identify better or adjunctive therapeutics. Future clinical trials with larger sample sizes, combined with immune, or omics testing, are necessary to clarify the indications for immune treatments and their mechanisms in PANS.

Firstly, we found that children with PANS exhibited baseline immune dysregulation compared to healthy controls, with predominantly downregulated immune pathways. However, NK cells exhibited upregulated immune pathways at baseline, suggesting that ‘immune dysregulation’ is a better description than ‘immune deficiency’ alone.

While our study focused on peripheral immune cells, microglia (resident central nervous system immune cells) dysregulation has also been implicated in PANS(8). We hypothesize that an impaired innate immune response, both peripherally and in the central nervous system, increases infection susceptibility, with recurrent infections and chronic inflammation triggering and sustaining symptoms in PANS.

Following IVIg treatment, the immune pathways that were downregulated at baseline were significantly upregulated compared to baseline. Previous single-cell RNA sequencing studies have also revealed cell-specific mechanistic actions of IVIg in Kawasaki disease, demonstrating that its effects are not uniform but vary according to cell type(9). IVIg has multiple immunomodulatory mechanisms of action, including saturation of Fc receptors, neutralization of idiotypic antibodies, complement inactivation, and cytokine inhibition. We found broad immune-modulating effects of IVIg across all peripheral blood cell types. However, its impact on peripheral-central immune crosstalk and central nervous system cells requires further investigation.

Secondly, we identified ribosomal dysregulation at baseline in children with PANS compared to healthy controls, which was reversed following IVIg treatment. The ribosomal machinery is increasingly recognised to play a role in NDDs(10). During times of cellular stress triggered by environmental factors, ribosome function is altered to conserve energy and promote stress adaptation. Epigenetic machinery, including chromatin remodelling, histone modifications, and stress-responsive transcription factors, play a crucial role in regulating ribosome function during stress by modulating gene expression and translation(11).

There is increasing support for the role of epigenetics, such as chromatin-related genes, in the pathogenesis of NDDs(12). Monogenic epigenetic disorders involving pathogenic DNA variants in chromatin-related genes such as *KMT2D, CDH7, HIST1HIE* have been described in children with infection provoked neuroregression or PANS-like presentations (13). Although children with PANS in this study did not have pathogenic DNA variations in chromatin-related or other epigenetic genes, it is plausible that epigenetic mechanisms play a role in PANS, potentially influenced by common gene variants plus environmental factors. Animal models suggest that maternal inflammatory factors during pregnancy, such as infection-mimics, autoimmune conditions, and obesity, can influence offspring brain and immune function, potentially through epigenetic mechanisms(14). The prenatal inflammatory environment is proposed to prime microglia epigenetically, heightening vulnerability to secondary insults later in life, leading to abnormal behaviour(14, 15). In this study, we observed a high incidence of maternal autoimmunity among our patients, consistent with previous findings in children with PANS (16). Additionally, we identified epigenetic effects, mainly histone and chromatin modifications, of IVIg on peripheral immune cells in PANS. In the NK cell histone methyltransferase activity pathway, histone-related genes, including *KMT, SETD*, and *NSD,* were generally upregulated at baseline in PANS pre-IVIg compared to controls, followed by downregulation after IVIg treatment. The epigenetic effects of IVIg and its downstream impact on immune and ribosomal function should be explored further.

This study shows the strength of single-cell RNA sequencing technology as a tool to discover biomarkers and explore therapeutic mechanisms of action in small cohorts such as this(17). Limitations of this study include the small sample size and the use of single-cell RNA sequencing on peripheral blood immune cells rather than brain tissue. However, we believe that peripheral blood RNA sequencing will transpire to be a useful biomarker that reflects gene regulation (epigenetics) in NDDs (5). Further functional immune testing is essential to confirm and better characterize the immune dysregulation that we identified through RNA sequencing. Although we identified changes in the expression of epigenetic genes, further targeted epigenetic analyses such as histone modification profiling, ATAC sequencing, or methylation analysis, will be essential to further explore the role of epigenetics in PANS and the effects of IVIg. Additionally, given the transient effect of IVIG observed in these patients, identifying alternative oral or dietary treatments with similar immune and epigenetic mechanisms of action could be useful.

## Conclusion

In five children with severe PANS who clinically responded to IVIg, we identified baseline abnormalities in epigenetic and ribosomal regulation, along with downstream immune dysfunction in PANS, and demonstrate that IVIg exerts both epigenetic and immune effects.

## Conflict of interest

There are no competing interests to declare.

## Data Availability

Anonymized data not published within this article will be made available by request from any qualified investigator.

## Acknowledgements

We are grateful for the study participants who contributed to this research. We would like to thank Australian Genome Research Facility for providing expertise in sequencing and bioinformatic support. We would like to thank PANS Australia, Stepping Stones Therapy for Children, and Dr Leila Masson for their kind donations for this study.

## Author contributions

VH: Data curation, Formal analysis, Investigation, Methodology, Roles/Writing-original draft; HN: Data curation, Formal analysis, Investigation, Roles/Writing- review & editing; BK: Data curation, Formal analysis, Investigation, Roles/Writing- review & editing; BG: Formal analysis, Investigation, Methodology, Roles/Writing- review & editing; XL: Formal analysis, Investigation, Methodology, Roles/Writing- review & editing; RD: Formal analysis, Investigation, Methodology, Roles/Writing- review & editing; JH: Data curation, Formal analysis, Investigation, Roles/Writing- review & editing; SM: Data curation, Formal analysis, Investigation, Roles/Writing- review & editing; SP: Conceptualization, Data Curation, Investigation, Methodology, Supervision, Writing- review & editing; RCD: Conceptualization, Data Curation, Investigation, Formal analysis, Funding acquisition, Methodology, Supervision, Writing- review & editing SP and RD share responsibilities as senior authors.

## Funding

Financial support for the study was granted by the Dale NHMRC Investigator Grant AP1193648, Petre Foundation, Cerebral Palsy Alliance, PANS Australia, PANDAS Network and International OCD Foundation. VXH was supported by ExxonMobil NUS Research Scholarship and National University Hospital Singapore (NUHS) Clinician-Scientist Program during the course of this study.

## Notes

### Competing Interest Statement

The authors have declared no competing interest.

### Author Declarations

Human Research Ethics Committee of Sydney Children Hospitals Network gave ethical approval for this work.

