## Supplementary Figures for "Intravenous immunoglobulin has epigenetic, ribosomal, and immune effects in Paediatric Acute-Onset Neuropsychiatric Syndrome"

### Slide 1
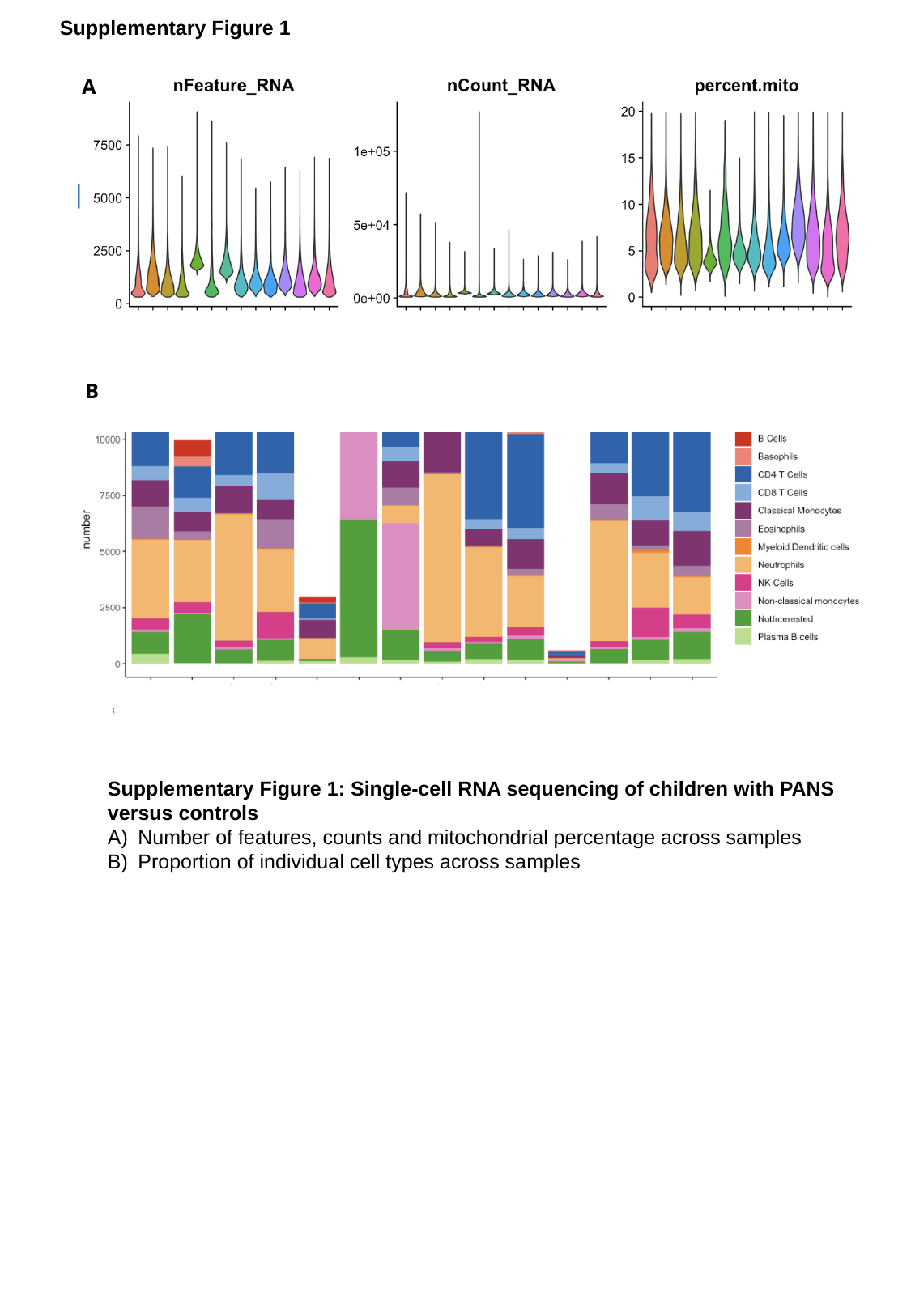

Supplementary Figure 1
A
B
Supplementary Figure 1: Single-cell RNA sequencing of children with PANS
versus controls
Number of features, counts and mitochondrial percentage across samples
Proportion of individual cell types across samples

### Slide 2
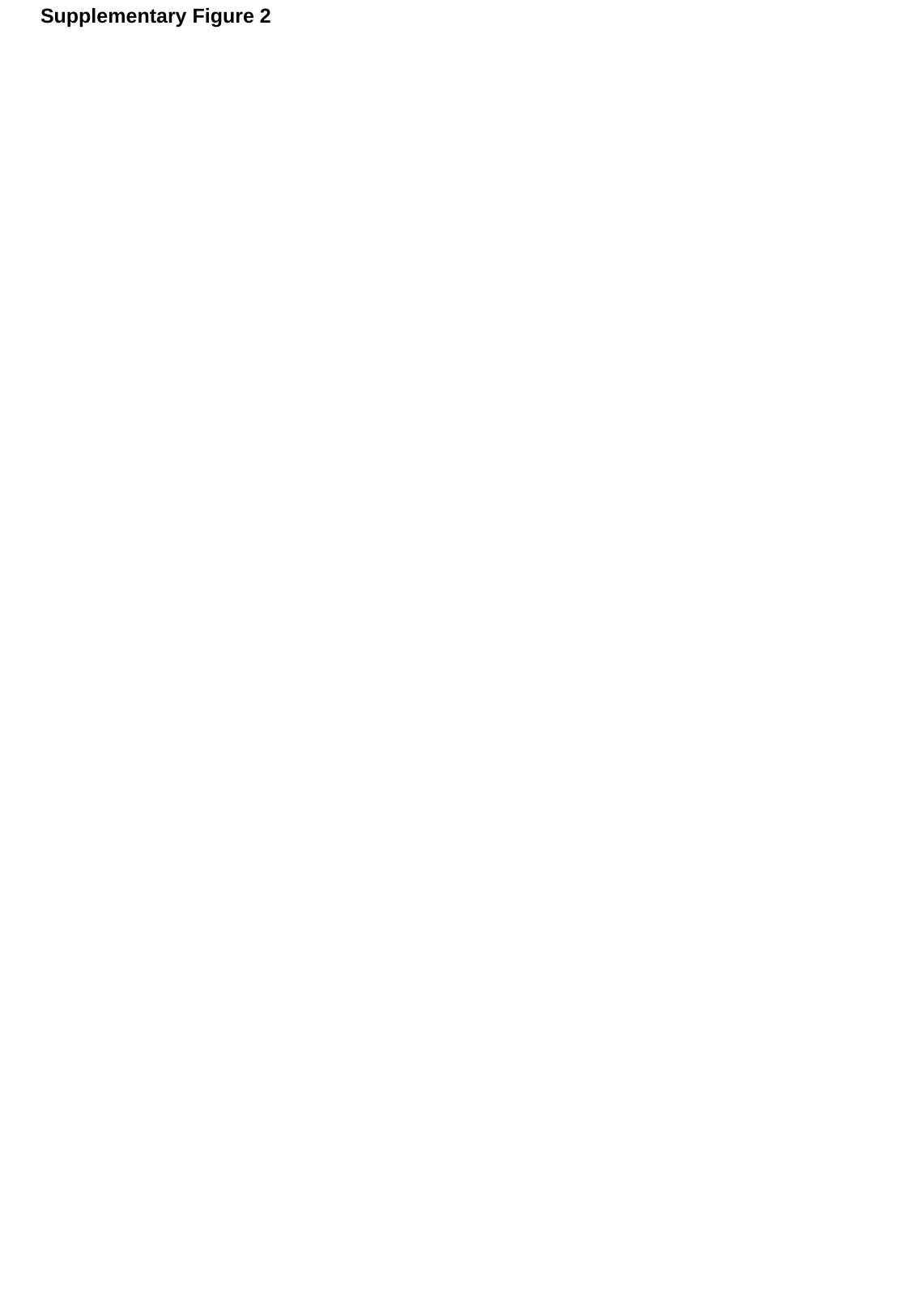

Supplementary Figure 2

### Slide 3
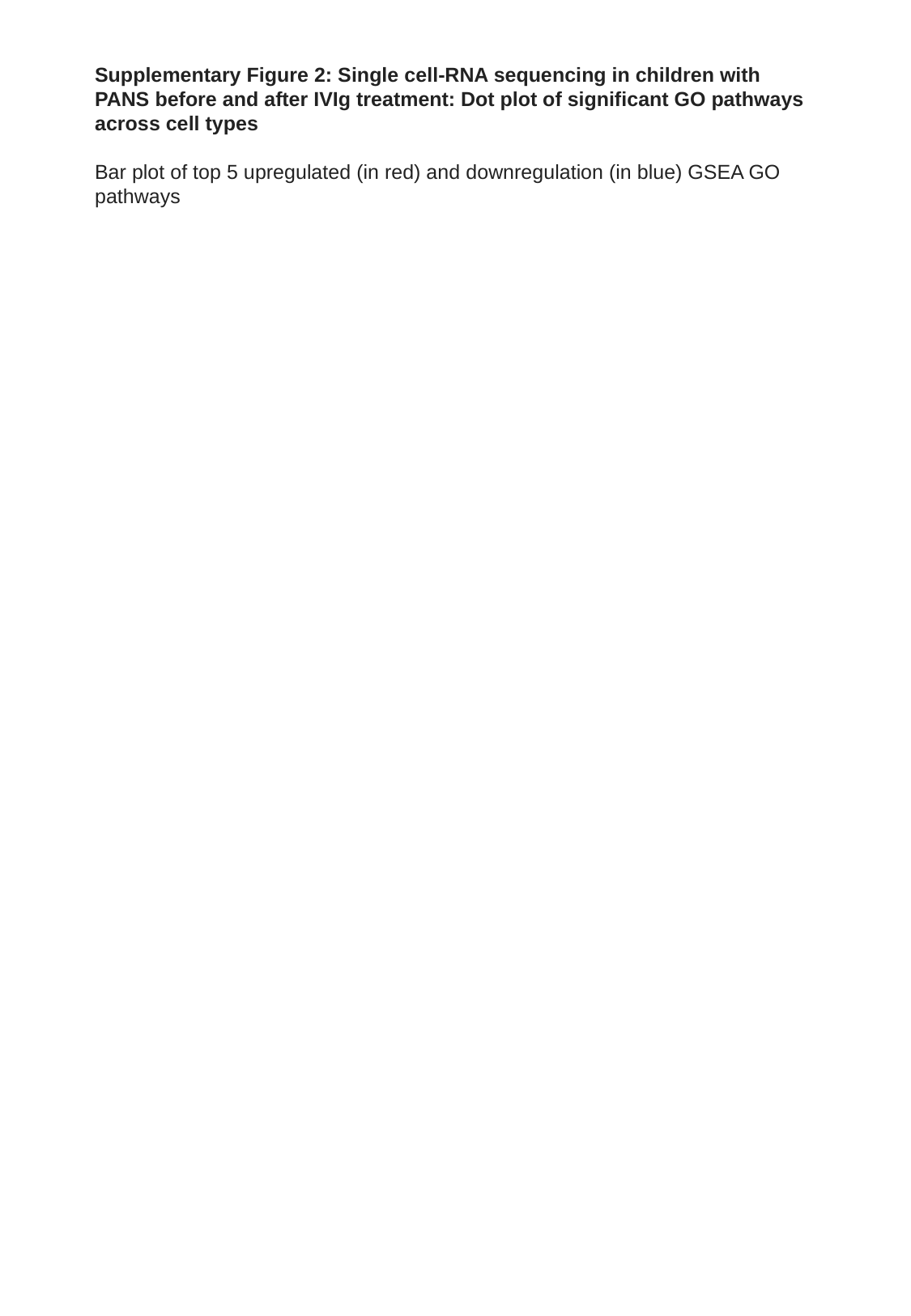

Supplementary Figure 2: Single cell-RNA sequencing in children with PANS before and after IVIg treatment: Dot plot of significant GO pathways across cell types
Bar plot of top 5 upregulated (in red) and downregulation (in blue) GSEA GO pathways
