## Supplementary Materials for "Intravenous immunoglobulin has epigenetic, ribosomal, and immune effects in Paediatric Acute-Onset Neuropsychiatric Syndrome"

**Supplementary Method- Single cell RNA sequencing**

*Single-cell blood RNA sequencing method*

Red blood cells were depleted from whole blood by immunomagnetic negative selection, using the EasySep™ RBC depletion reagent protocol (catalog #18000), preserving all leukocyte populations, including granulocytes. 14 HIVE devices (CLX version) (individual samples from 5 patients (pre- and post-IVIg treatment) versus 4 controls) were each loaded with approximately 30,000 cells in 1 mL of DPBS + 1% FBS, followed by 3 mL of cell media (DPBS + 1% FBS). The leukocytes were loaded into the HIVE system within an hour of blood sampling from all individuals to minimize neutrophil activation. Single cells settled into picowells within the HIVE devices, which contained 3' transcript-capture beads. HIVEs were centrifuged at 30 x g for 3 minutes to ensure cell settling. Following media removal, HIVEs were washed twice with 2 mL of sample wash solution. After removing the wash solution, 2 mL of cell preservation solution was added, and the cell-loaded HIVEs were frozen at -80°C. Frozen devices were then shipped to the Australian Genome Research Facility Ltd (AGRF Ltd, Westmead) for single-cell next-generation sequencing (NGS) library preparation, according to the manufacturer's protocol ([Honeycomb Biotechnologies, Inc. USA_Sample Capture protocol](https://honeycombbio.zendesk.com/hc/en-us/articles/15173728631707-HIVE-CLX-Sample-Capture-User-Protocol)).

Subsequently, HIVE devices were sealed with a semi-permeable membrane, enabling on-device lysis and hybridization. Beads with captured transcripts were extracted from the HIVE device by centrifugation, and subsequent library preparation steps were performed in a 96-well plate format ([Honeycomb Biotechnologies, Inc. USA: Transcriptome Recovery and Library Preparation protocol](https://honeycombbio.zendesk.com/hc/en-us/articles/15173667166491-HIVE-CLX-Transcriptome-Recovery-Library-Preparation-User-Protocol)). Library size distribution and quality were assessed using a TapeStation 2200 platform with a D5000 ScreenTape System (Agilent Technologies, Santa Clara, CA, USA). The concentration of final pooled libraries was determined by qPCR. Final HIVE scRNA-seq libraries were sequenced on an Illumina NovaSeq X sequencer (AGRF Ltd, Melbourne) using kit-specific primers.

*Single-cell RNA sequencing bioinformatic analysis*

Cells with a high mitochondrial transcript ratio (>0.15) were excluded. Experiments were integrated using the *FindIntegrationAnchors* function in *Seurat* then immune cell types were assigned using *scPred*. Merged data were then split by cell type and separately normalized, scaled, and integrated between patients using *harmony*, then UMAP (uniform manifold approximation and *projection)* projections were made using the first 30 dimensions. Differentially expressed genes were identified using *FindMarkers.*

*Gene set enrichment (GSEA) pathway analyses*

The genes were ranked based on their logFC. Enriched gene sets were identified based on a running sum statistic (normalized enrichment score (NES)) and statistical significance, based on the false discovery rate (FDR). Significant GSEA GO pathways (FDR <0.05) were further simplified using the *simplify* function in clusterProfiler.

*Bar/dot and connectivity network plots*

Bar and dot plots of GSEA gene ontology (GO) results were plotted using *ggplot2* package. The top 5 up-regulated and down-regulated GSEA GO post-simplified pathways, for each cell type were selected as representative GO terms. For the dot plot of 7 cell types, only the GO terms present in more than 3 cell types in the pre-IVIg vs control comparisons were mapped.

Connectivity network (CNET) plots were created using *enrichplot* package where the enriched pathways are represented by their respective colors. To further evaluate themes within pathways, gene subclusters were created based on most significant Gene Ontology pathways per gene set and CNET plots were created. The enriched pathways were represented by their respective colors and corresponding gene associated with the pathway. Heatmaps of individual genes in GO pathway “histone methyltransferase activity” in NK cells were made using the *pheatmap* package.
